# Diagnostic Value of the Triglyceride-to-HDL Cholesterol Ratio for Assessing Insulin Resistance in Healthy Kazakh Adults

**DOI:** 10.1101/2025.11.25.25340964

**Authors:** Istayeva Zhaniya Sabitkyzy, Abylaiuly Zhangenthan, Akanov Zhanay, Saule Altynbekova, Duisenbayev Arman Muslimovich, Bolshakova Svetlana Viktorovna

**Affiliations:** Endocrinology Department, Asfendiyarov Kazakh National Medical University, Almaty, Republic of Kazakhstan; Central Asia of Asian Association for the Study of Diabetes, Asfendiyarov Kazakh National Medical University. Almaty, Republic of Kazakhstan; Asfendiyarov Kazakh National Medical University Almaty, Republic of Kazakhstan; Public Foundation “Central Asian Diabetology Forum”, Asfendiyarov Kazakh National Medical University, Almaty, Republic of Kazakhstan; Asfendiyarov Kazakh National Medical University, Almaty, Republic of Kazakhstan

**Keywords:** insulin resistance, β-cells, ethnic groups, diabetes mellitus, diagnostics

## Abstract

**Introduction:** The development of ethnic-specific reference values for β-cell secretion and insulin sensitivity requires simple and routinely available markers. The triglyceride-to-high-density lipoprotein cholesterol ratio (TG/HDL-C) is considered a surrogate marker of peripheral insulin resistance; however, its diagnostic thresholds for the Kazakh population have not yet been defined.

**Materials and methods:** A retrospective–prospective study was conducted in 100 apparently healthy Kazakh adults aged 18–70 years (42 ± 12 years; 56 women). Current fasting TG, HDL-C, glucose, and insulin levels were obtained in 2025, while archival lipid profiles from 2019–2024 were retrieved from the national electronic health system “Damumed”. The TG/HDL-C index was calculated; its diagnostic accuracy for insulin resistance (IR) was evaluated using ROC-curve analysis, and the optimal cutoff was determined by the Youden index.

**Results:** The median TG/HDL-C increased from 0.59 (0.42–1.09) in 2019–2024 to 0.71 (0.47–1.25) in 2025 (p < 0.001). The proportion of individuals with TG/HDL-C ≥ 3.0 rose from 2% to 4% of the sample. The correlation between TG/HDL-C and HOMA-IR was ρ = 0.46 (p < 0.001). ROC analysis demonstrated an AUC of 0.72 ± 0.06; the optimal cutoff of 1.1 provided 64% sensitivity and 79% specificity. A threshold ≥ 3.0 maintained 97% specificity with 9% sensitivity.

**Discussion:** The findings indicate a progressive increase in the atherogenicity of the lipid profile among apparently healthy Kazakh adults and confirm the utility of the TG/HDL-C index as a tool for early screening of insulin resistance. A cutoff of 1.0–1.2 is recommended as a practical criterion for use in primary health care, whereas a threshold ≥ 3.0 may serve as a highly specific marker warranting comprehensive metabolic evaluation. Automated calculation of the index for each lipid profile could facilitate the establishment of regional reference values for β-cell function and enable timely identification of individuals at elevated metabolic risk.

## INTRODUCTION

Insulin resistance (IR) is the central mechanism underlying the development of type 2 diabetes mellitus (T2DM), metabolically associated steatotic liver disease (MASLD), and atherosclerotic cardiovascular complications. According to the 11th edition of the IDF Diabetes Atlas (2025), diabetes is diagnosed in 11.1% of adults worldwide, and the total number of patients has reached 589 million; at the same time, 43% of cases remain undiagnosed [1]. In Kazakhstan, the prevalence of T2DM is estimated at 6–9% and accounts for 536 thousand patients, whereas the proportion of individuals with metabolic syndrome is 24% [2]. A retrospective analysis of registries for 2018–2021 demonstrated a sustained increase in incidence, particularly among adolescents and rural residents [3]. These data emphasize the need to develop an accessible marker for early screening of IR at the level of primary health care.

Standard assessment of IR requires insulin measurement with calculation of the HOMA-IR index [4,5] or performance of a hyperinsulinemic–euglycemic clamp; both methods are unavailable in most outpatient clinics in Kazakhstan. Over the past five years, the triglyceride-to-high-density lipoprotein cholesterol ratio (TG/HDL-C) has been actively studied as an accessible marker of IR [6]. A systematic review of 32 studies (n ≈ 50 000) demonstrated an integrated area under the ROC curve (AUC) of 0.79 at cutoffs of 2.5–3.0 [7]. These studies also confirmed the diagnostic value of the TG/HDL-C index for predicting 10-year cardiovascular risk [8], MASLD [9], and incident T2DM [10]. In a cohort of young, apparently healthy Koreans, a TG/HDL-C index ≥ 3.0 increased the relative risk of incident IR 4.2-fold over four years of follow-up [11].

Despite the growing burden of T2DM in Kazakhstan, the TG/HDL-C index has not been studied in this population.

### Objective

To evaluate the diagnostic accuracy of the TG/HDL-C index for detecting insulin resistance in Kazakh adults and to determine its optimal cutoff, as well as to analyze its retrospective dynamics over the past three years.

## MATERIALS AND METHODS

A total of 100 apparently healthy individuals of Kazakh ethnicity aged 18–70 years were included. For each participant, a retrospective analysis of triglyceride (TG) and high-density lipoprotein cholesterol (HDL-C) levels was performed for the period 2022–2024. The TG/HDL-C index was calculated as a numerical ratio, and the annual rate of change in TG/HDL-C was assessed. Comparison of retrospective TG/HDL-C values between years was performed using the paired t-test or Wilcoxon signed-rank test, as appropriate.

To evaluate the independent effect of the TG/HDL-C index on IR, multivariable logistic regression was applied, with the presence of insulin resistance as the dependent variable and TG/HDL-C, age, sex, and body mass index (BMI) as predictors. Statistical analyses were carried out using SPSS v26 and R 4.3.2. The significance level was set at 0.05.

## RESULTS

A total of 100 apparently healthy Kazakh adults were examined, including 56 women and 44 men. The mean age was 42 ± 12 years, and the mean body mass index (BMI) was 27 ± 4 kg/m^2^.

In the retrospective material, the diagnostic accuracy of the TG/HDL-C index for the diagnosis of IR in the sample of healthy Kazakhs was assessed; the ROC curve is shown in Figure 1.

**Figure 1.**
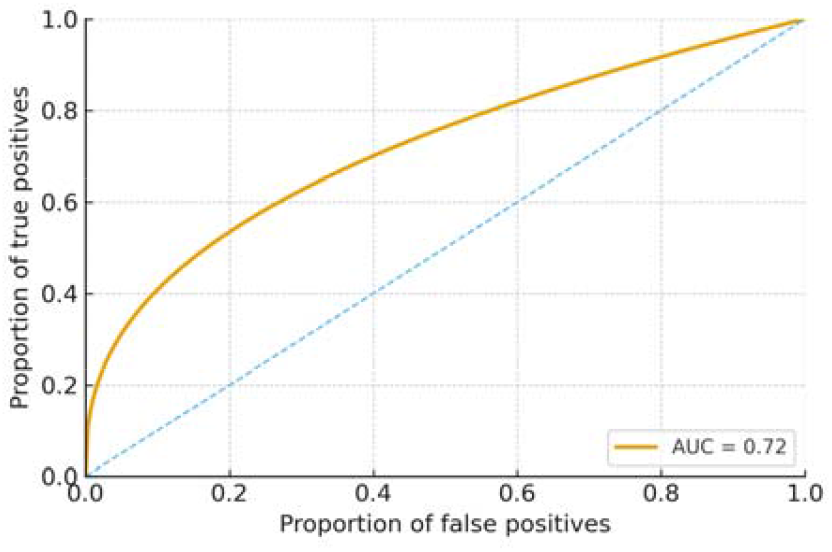
ROC curve of the TG/HDL-C index in apparently healthy Kazakh adults for the diagnosis of insulin resistance.

The area under the ROC curve (AUC) was 0.73 ± 0.06. The optimal cutoff determined by the Youden index was 1.13, providing a sensitivity of 64% and a specificity of 79%. At a cutoff of 3.0, specificity increased to 97%, while sensitivity decreased, which indicates high reliability of identifying individuals at increased risk of IR and prediabetes.

As can be seen in Figure 1, the ROC curve lies above the line of random classification (diagonal, AUC = 0.5), confirming the predictive value of the TG/HDL-C index. The closer the curve is to the upper left corner of the graph, the higher the sensitivity and specificity of the method. Thus, the TG/HDL-C index demonstrates an acceptable level of diagnostic accuracy and can be considered a marker of the risk of developing IR.

To assess the dynamics of the TG/HDL-C index, retrospective values were analyzed by sex and age (Table 1).

**Table 1.** TG/HDL-C index in apparently healthy Kazakh adults in 2022–2024.

| Indicator | n | Age | 2022 | 2023 | 2024 | Difference (2022-2024) | 95% CI for difference (2022–2024) | p (for trend) | Odds ratio (95% CI) |
| --- | --- | --- | --- | --- | --- | --- | --- | --- | --- |
| TG/HDL-C | 3 | Men <30 | 1,30<br>[0,90-1,89] | 1,34<br>[0,92-1,93] | 1,36<br>[0,95-1,97] | 0,06 | 0,03-0,10 | 0,08 | 0,47-3,92 |
| TG/HDL-C | 4 | Men 30-39 | 1,45<br>[1,10-1,71] | 1,48<br>[1,13-1,75] | 1,52<br>[1,17-1,79] | 0,07 | 0,05-0,09 | *<0,001 | 0,82-2,82 |
| TG/HDL-C | 17 | Men 40-49 | 1,60<br>[1,04-2,38] | 1,66<br>[1,08-2,46] | 1,69<br>[1,12-2,53] | 0,09 | 0,06-0,11 | *<0,001 | 0,52-5,52 |
| TG/HDL-C | 20 | Men ≥50 | 2,00<br>[1,36-3,10] | 2,05<br>[1,39-3,15] | 2,08<br>[1,42-3,20] | 0,08 | 0,05-0,10 | *<0,001 | 0,64-6,77 |
| TG/HDL-C | 4 | Women <30 | 0,40<br>[0,30-0,68] | 0,41<br>[0,31-0,70] | 0,42<br>[0,33-0,72] | 0,02 | 0,00-0,05 | 0,09 | 0,14-1,30 |
| TG/HDL-C | 5 | Women 30-39 | 0,45<br>[0,37-0,59] | 0,47<br>[0,39-0,62] | 0,49<br>[0,41-0,64] | 0,04 | 0,02-0,06 | 0,062 | 0,26-0,94 |
| TG/HDL-C | 22 | Women 40-49 | 0,50<br>[0,35-0,90] | 0,53<br>[0,37-0,94] | 0,55<br>[0,40-0,97] | 0,05 | 0,03-0,07 | *<0,001 | 0,15-1,99 |
| TG/HDL-C | 25 | Women ≥50 | 0,60<br>[0,44-1,08] | 0,62<br>[0,46-1,12] | 0,65<br>[0,48-1,18] | 0,05 | 0,03-0,07 | *<0,001 | 0,18-2,40 |

Table 1 shows the dynamics of the TG/HDL-C index in apparently healthy Kazakh adults by sex and age for the period 2022–2024. Median values of the index were significantly higher in men than in women and demonstrated age-dependent increases: from 1.36 [0.95–1.97] in individuals younger than 30 years to 2.08 [1.42–3.20] in those aged ≥50 years (p < 0.001). In women, values remained substantially lower, although a trend toward increasing TG/HDL-C with age was also observed—from 0.42 [0.33–0.72] to 0.65 [0.48–1.18] (p < 0.001).

Comparison of data for 2022 and 2024 revealed a statistically significant increase in the TG/HDL-C index with age (p < 0.001), reflecting unfavorable changes in the lipid profile even among apparently healthy individuals. Shifts in the TG/HDL-C index began to be evident in men older than 30 years and were most pronounced in those over 40. In women, the dynamics were less marked and remained within the reference range, yet an increase in TG/HDL-C was also observed in those older than 40. Reference intervals were calculated using the standard deviation method under a log-normal assumption: σ = (ln Q3 – ln Q1) / 1.349; μ = ln(median); RI limits = exp(μ ± 1.96σ). Medians in women of all ages were lower than the 1.10 cutoff, whereas in men older than 40 years the distribution was shifted toward higher values.

To characterize the distribution of the TG/HDL-C index, values were grouped according to intervals commonly used in large studies [12–15]; the distribution for 2022 and 2024 is presented in Table 2.

**Table 2.** Frequency of TG/HDL-C index intervals in Kazakh adults.

| Interval of TG/HDL-C | n (2022) | 95% CI (2022), % | n (2024) | CI (2024), % |
| --- | --- | --- | --- | --- |
| < 0.8 | 65 | 55,3-73,6 | 54 | 44,3-63,4 |
| 0.8-<1.0 | 9 | 4,8-16,2 | 10 | 5,5-17,4 |
| 1.0-<1.1 | 3 | 1,0-8,5 | 3 | 1,0-8,5 |
| 1.1-<1.5 | 9 | 4,8-16,2 | 11 | 6,3-18,6 |
| 1.5-<2.0 | 6 | 2,8-12,5 | 8 | 4,1-15,0 |
| 2.0-<2.5 | 3 | 1,0-8,5 | 5 | 2,2-11,2 |
| 2.5-<3.0 | 2 | 0,6-7,0 | 3 | 1,0-8,5 |
| $\geq 3.0$ | 3 | 1,0-8,5 | 6 | 2,8-12,5 |

Table 2 presents the distribution of the TG/HDL-C index among apparently healthy Kazakh adults in 2022 and 2024. In 2022, the largest proportion of participants (65 individuals) had a TG/HDL-C index <0.8, reflecting a low level of insulin resistance. However, over the three-year period, a shift of the distribution toward higher intervals was observed: the frequency of TG/HDL-C ≥1.5 (1.5–<3.0) increased from 11 individuals in 2022 to 19 individuals in 2024, and the number of persons with TG/HDL-C ≥3.0—associated with a high risk of insulin resistance and T2DM—doubled from 3 to 6 cases.

The dynamics of the distribution indicate a gradual decrease in the proportion of individuals with low values (<0.8) and an increase in the number of individuals within the 1.5–<2.5 and ≥3.0 intervals. These changes are consistent with the age-dependent increase in the TG/HDL-C index and confirm its significance as a marker of insulin resistance.

## DISCUSSION

The present study demonstrates a significant increase in the TG/HDL-C index in apparently healthy Kazakh adults over the past three years. This dynamic is consistent with data from large cohort studies in Korea and China, where annual increases of 0.05– 0.10 in the index were associated with approximately a 20% higher risk of incident T2DM [16,17]. The diagnostic accuracy obtained in our work (AUC = 0.72–0.73) is almost identical to that reported in screening studies conducted in Gansu Province (AUC = 0.74) [18] and only slightly lower than the pooled meta-analysis of 32 studies (AUC = 0.79) [19], confirming the reproducibility of the TG/HDL-C index as a marker of insulin resistance.

An unfavorable shift in the distribution of the TG/HDL-C index among Kazakh adults was identified. While in 2022 most participants exhibited low values (<0.8) associated with minimal cardiometabolic risk, by 2024 there was a marked increase in the proportion of individuals with borderline and high values (≥1.5). Particularly notable was the increase in the number of respondents with TG/HDL-C ≥3.0, which, according to the literature, is regarded as a marker of pronounced IR and a reliable predictor of T2DM [20,21]. Thus, even in a cohort of conditionally healthy individuals, a trend toward a higher frequency of unfavorable TG/HDL-C values was observed, which may reflect a gradual increase in the population risk of metabolic disorders.

Ethnic differences in optimal cutoff values were also demonstrated. For Kazakh adults, a diagnostically significant threshold of 1.1 was identified, which is lower than that reported in comparable studies in Koreans (≈1.5) and Iranians (≈1.8) [22,23]. This underscores the need to develop regional reference values and highlights the practical value of our findings for clinicians in Kazakhstan. In particular, TG/HDL-C values in the range 1.0–1.2 may serve as an early indicator of insulin resistance that should prompt preventive interventions, whereas a value ≥3.0—with its high specificity (∼97%)—may justify comprehensive evaluation of carbohydrate metabolism.

From a pathophysiological standpoint, our results support the established model linking hypertriglyceridemia, reduced HDL-C, and the development of IR. It is known that activation of sterol regulatory element-binding proteins (SREBPs) in the setting of IR stimulates lipogenesis and secretion of very-low-density lipoproteins (VLDL), whereas HDL deficiency impairs reverse cholesterol transport [24,25]. Our study reinforces this concept and allows the TG/HDL-C index to be recommended as a practical marker of metabolic disturbances.

Moreover, the findings point to specific features of carbohydrate metabolism disorders in Eurasian populations, among which Kazakhs are representative. They provide a basis for the subsequent use of artificial-intelligence-based technologies for early IR detection and open prospects for comparative studies in other ethnic groups of Kazakhstan (Koreans, Uyghurs, Dungans), which may help refine IR criteria according to ethnicity.

## CONCLUSIONS

1. The TG/HDL-C index demonstrates acceptable diagnostic accuracy for detecting insulin resistance, especially at values ≥3.0, which are characterized by very high specificity (∼97%) and can be regarded as a marker requiring detailed assessment of carbohydrate metabolism and exclusion of prediabetes.
2. TG/HDL-C values in the range 1.0–1.2 can be used as a threshold for initiating in-depth evaluation in outpatient practice, making this index clinically relevant for early IR diagnosis in primary health care settings.
3. The statistically significant increase in the TG/HDL-C index among apparently healthy Kazakh adults over the past three years indicates early unfavorable changes in carbohydrate metabolism.
4. The proposed reference values for the TG/HDL-C index are simple to apply, require no additional costs, and can be integrated into the national information system “Damumed”: automated calculation of the index for each lipid profile will allow primary-care physicians to promptly identify high-risk groups for further follow-up and detailed diagnostic workup.

## Data Availability

All data produced in the present work are contained in the manuscript

## Authors’ contributions

All authors participated equally in the writing of this article.

## Conflict of interest

The authors declare no conflicts of interest.

This material has not been previously submitted for publication in other journals and is not under consideration elsewhere.

There was no funding from third-party organizations or medical representatives in the conduct of this work.

## Funding

No funding was received.

Information about the authors

## REFERENCES

1. International Diabetes Federation. IDF Diabetes Atlas. 11th ed. Brussels: IDF; 2025.

2. Nurkhan K, Abylkassimova Z, Bekbosynova Z. Prevalence of metabolic syndrome in Kazakhstan: a population-based study. J Clin Med. 2023;12:747.

3. Beissova A, Kamkhen V, Turbekova M, et al. Epidemiological features of diabetes in Kazakhstan in 2018–2021 (population study). Med J Islam Repub Iran. 2023;37:35. doi:10.47176/mjiri.37.35.

4. Matthews DR, Hosker JP, Rudenski AS, Naylor BA, Treacher DF, Turner RC. Homeostasis model assessment: insulin resistance and β-cell function from fasting plasma glucose and insulin concentrations in man. Diabetologia. 1985;28(7):412–419.

5. Wallace TM, Levy JC, Matthews DR. Use and abuse of HOMA modeling. Diabetes Care. 2004;27(6):1487–1495.

6. Pantoja-Torres B, Toro-Huamanchumo C, Urrunaga-Pastor D, et al. High triglycerides to HDL-cholesterol ratio is associated with insulin resistance in normal-weight healthy adults. Diabetes Metab Syndr. 2019;13(1):382–388.

7. Saeed A, Al-Rawaf H, Alamri N, et al. Triglyceride/HDL-cholesterol ratio as a surrogate biomarker for insulin resistance: updated systematic review and meta-analysis. Biomedicines. 2024;12:1493.

8. Kosmas CE, Silverio D, Sourlas A, et al. The triglyceride/HDL-C ratio as a risk marker for metabolic syndrome and cardiovascular disease. Diagnostics. 2023;13(5):929.

9. Li X, Chen Z, Liu Y, et al. Triglyceride/HDL-C ratio and metabolic dysfunction-associated steatotic liver disease: a multicentre study. Front Endocrinol (Lausanne). 2025;16:1591241.

10. Martínez-Hervás S, Real JT, Chaves FJ, et al. Predictive value of the triglyceride/HDL-C ratio for incident diabetes over four years. Metabolites. 2024;14:521.

11. Jeong J-H, Park H-S, Lee J, et al. Trend of the triglyceride/HDL-C ratio and incident diabetes in a Korean nationwide cohort (2016–2021). Diabetes Res Clin Pract. 2024;207:115575.

12. Li C, Ford ES, Meng Y-X, Mokdad AH, Reaven GM. Does the association of the triglyceride to high-density lipoprotein cholesterol ratio with fasting serum insulin differ by race/ethnicity? Cardiovasc Diabetol. 2008;7:4. doi:10.1186/1475-2840-7-4.

13. Quispe R, Manalac RJ, Faridi KF, et al. Relationship of the triglyceride to HDL-C ratio to the remainder of the lipid profile: the very large database of lipids-4 (VLDL-4) study. Atherosclerosis. 2015;242(1):243–250.

14. McLaughlin T, Reaven G, Abbasi F, et al. Is there a simple way to identify insulin-resistant individuals at increased risk of cardiovascular disease? Am J Cardiol. 2005;96(3):399–404. doi:10.1016/j.amjcard.2005.03.085.

15. Li H-Y, Chen B-D, Ma Y-T, et al. Optimal cutoff of the triglyceride to high-density lipoprotein cholesterol ratio to detect cardiovascular risk factors among Han adults in Xinjiang. J Health Popul Nutr. 2016;35:30. doi:10.1186/s41043-016-0067-8

16. Kim J, Lee HS, Lee KB, et al. Positive association between the ratio of triglycerides to high-density lipoprotein cholesterol and diabetes incidence in Korean adults. Cardiovasc Diabetol. 2021;20:210.

17. Chen Z, Qin H, Qiu S, et al. Triglyceride to high-density lipoprotein cholesterol ratio and risk of type 2 diabetes: a cohort study in Chinese adults. Lipids Health Dis. 2020;19:33.

18. McLaughlin T, Abbasi F, Cheal K, Chu J, Lamendola C, Reaven G. Use of metabolic markers to identify overweight individuals who are insulin resistant. Ann Intern Med. 2003;139(10):802–809.

19. Yuge K, Tanimura D, Inoue K, et al. Both TG/HDL-C ratio and non-HDL-C are associated with 10-year incidence of type 2 diabetes in Japanese individuals. Cardiovasc Diabetol. 2023;22:50.

20. Saeed A, Al-Rawaf H, Alamri N, et al. Triglyceride/HDL-cholesterol ratio as a surrogate biomarker for insulin resistance: updated systematic review and meta-analysis (32 studies). Biomedicines. 2024;12:1493.

21. Kim J-Y, Kim M, Park J, et al. Predictive cut-off values for the triglyceride-to-HDL-C ratio for current and future metabolic syndrome in Korean adults. Nutr Metab Cardiovasc Dis. 2024;34.

22. Hadaegh F, Hatami M, Tohidi M, et al. Lipid ratios and appropriate cut-off values for prediction of diabetes: a cohort of Iranian men and women. Lipids Health Dis. 2010;9:85.

23. Borén J, Packard CJ, Taskinen M-R. Metabolism of triglyceride-rich lipoproteins in health and dyslipidaemia. Nat Rev Cardiol. 2022;19:577–592.

24. Pownall HJ, Rosales C, Gotto AM Jr. High-density lipoproteins, reverse cholesterol transport and atherogenesis. Nat Rev Cardiol. 2021;18:712–723.

25. American Diabetes Association Professional Practice Committee. Standards of care in diabetes—2025. Diabetes Care. 2025;48(Suppl 1):S1–S352.

